# IMPACT OF UNIVERSITY RE-OPENING ON TOTAL COMMUNITY COVID-19 BURDEN

**DOI:** 10.1101/2020.09.18.20197467

**Authors:** Lauren E. Cipriano, Wael M. R. Haddara, Gregory S. Zaric, Eva A. Enns

## Abstract

**Purpose:** Post-secondary students have higher than average contacts than the general population due to congregate living, use of public transit, high-density academic and social activities, and employment in the services sector. We evaluated the impact of a large student population returning to a mid-sized city currently experiencing a low rate of COVID-19 on community health outcomes. We consider whether targeted routine or one-time screening in this population can mitigate community COVID-19 impacts.

**Methods:** We developed a dynamic transmission model of COVID-19 subdivided into three interacting populations: general population, university students, and long-term care residents. We parameterized the model using the medical literature and expert opinion. We calibrated the model to the observed outcomes in a mid-sized Canadian city between March 1 and August 15, 2020 prior to the arrival of a relatively large post-secondary student population. We evaluated the impact of the student population (20,000 people arriving on September 1) on cumulative COVID-19 infections over the fall semester, the timing of peak infections, the timing and peak level of critical care occupancy, and the timing of re-engaged social and economic restrictions. We consider multiple scenarios with different student and general population COVID-19 prevention behaviours as well as different COVID-19 screening strategies in students.

**Results:** In a city with low levels of COVID-19 activity, the return of a relatively large student population substantially increases the total number of COVID-19 infections in the community. In a scenario in which students immediately engage in a 24% contact reduction compared to pre-COVID levels, the total number of infections in the community increases by 87% (from 3,900 without the students to 7,299 infections with the students), with 71% of the incremental infections occurring in the general population, causing social and economic restrictions to be re-engaged 3 weeks earlier and an incremental 17 COVID-19 deaths. Scenarios in which students have an initial, short-term increase in contacts with other students before engaging in contact reduction behaviours can increase infections in the community by 150% or more. In such scenarios, screening asymptomatic students every 5 days reduces the number of infections attributable to the introduction of the university student population by 42% and delays the re-engagement of social and economic restrictions by 1 week. Compared to screening every 5 days, one-time mass screening of students prevents fewer infections, but is highly efficient in terms of infections prevented per screening test performed.

**Discussion:** University students are highly inter-connected with the city communities in which they live and go to school, and they have a higher number of contacts than the general population. High density living environments, enthusiasm for the new school year, and relatively high rates of asymptomatic presentation may decrease their self-protective behaviours and contribute to increased community transmission of COVID-19 affecting at-risk members of the city community. Screening targeted at this population provides significant public health benefits to the community through averted infections, critical care admissions, and COVID-19 deaths.

## INTRODUCTION

The COVID-19 pandemic presents a substantial public health challenge for local, national, and international communities because the virus is highly transmissible,^1-3^ including prior to symptom onset,^4,5^ and infections initially present with a wide range of non-specific and sometimes mild symptoms.^6,7^ The relatively high rate of hospitalization and need for critical care among severe cases can quickly overwhelm community health care resources and result in substantial mortality.^8,9^

Many communities initially responded to COVID-19 with school closures and stay-at-home orders, which included closure of university campuses and conversion of all in-person instruction to online formats. Over the summer, universities began announcing plans for the fall term. Some universities opted to operate fully online for the fall.^10^ Others announced plans to partially or fully re-open campus and welcome students back for in-person instruction with COVID-19 mitigation strategies in place. These strategies included polices around mask wearing, limiting large gatherings, access to COVID-19 testing, reduced dormitory occupancy, and accommodations for isolating and quarantining students.^11^ While universities have the autonomy to make decisions about the level of on-campus activities offered to their students, their decisions have implications for the communities in which these campuses are located. University students live, work, and socialize both on and off campus, resulting in significant potential for on-campus COVID-19 outbreaks to spill over into the community and vice versa.^12^

Infectious diseases can spread rapidly through a university campus, as evidenced by outbreaks of serogroup B meningococcal disease, mumps,^13^ and the novel H1N1 influenza virus that emerged in 2009.^12,14-16^ Surges of COVID-19 cases have already been observed on the first campuses to re-open this fall,^17^ prompting many universities to abruptly change their fall semester plans.^18^ After 177 students tested positive for the novel coronavirus linked to at least four separate clusters, the University of North Carolina abruptly moved all undergraduate classes online after only a week of in-person instruction.^19^ High-density housing, including multiple roommates and shared facilities like bathrooms, as well as high levels of social activity puts the university population at particular risk for infectious disease transmission.^15^ Furthermore, past experience with the 2009 H1N1 pandemic indicates compliance with recommended public health precautions may be sub-optimal; despite public health guidance not to attend classes and other activities while ill during the H1N1 outbreak at a US university, half of students indicated interacting with a symptomatic individual in a classroom and nearly one-quarter indicated interacting with a symptomatic individual at a party or social activity.^15^

Given the unique features of university communities, several studies have modeled COVID-19 transmission dynamics specifically on university campuses and evaluated potential mitigation strategies.^18,20-24^ These studies used mathematical models of viral transmission dynamics, tailored to reflect a university context, in order to evaluate testing and contact tracing strategies, largely focusing on the question of how frequently to test asymptomatic students. All analyses concluded that frequent testing (sometimes multiple times a week) would be needed to contain COVID-19 outbreaks on campus. High frequency testing has been adopted by a number of public and private universities including, for example, the University of Illinois (twice per week by saliva testing^25^), Colby College (twice per week by nasopharyngeal swab^26^), Cornell University (twice weekly for students and faculty with student contact by self-collected anterior nares sampling^27^), and Harvard University (one to three days a week for students, staff, and faculty by saliva testing with frequency depending on types of on-campus activity^28^). However, most universities do not have the resources or infrastructure to support high-frequency testing.^26^

Furthermore, while some of these studies did include infections among students arising from off-campus community contact, these studies did not consider the impact of university student infections and university administration prevention and management decisions on the broader community in which that campus is situated. University students themselves may be at lower risk of severe COVID-19 disease due to age, but high infection rates on campus may spill over into the broader community, whose members are at higher risk for adverse COVID-19 outcomes. It is therefore important to quantify the expected impact of the arrival of a relatively large number of university students on the broader community in terms of incremental infections, hospitalizations, and COVID-19 mortality. Through the many interactions between the student population and the city in which they reside, COVID-19 mitigation policies targeted at university student communities and adopted by university leaders may have substantial public health implications for those in the surrounding community.

We developed a dynamic transmission model of COVID-19 to estimate the health impacts and health care resource demand in a COVID-19 outbreak in a representative mid-sized city with a relatively large destination college campus. We assumed a city experiencing a low level of COVID-19 activity going into the semester and explore how the on-campus arrival of the student population impacts COVID-19 health outcomes in the community. We consider different assumptions about student behaviours related to physical distancing and mask wearing, as well as the mitigating effects of targeted routine and one-time COVID-19 screening in the university population.

## METHODS

### OVERVIEW

We developed a dynamic compartmental model to simulate infection dynamics and health resource use of a representative mid-sized city with a population of 500,000 going into fall after experiencing low rates of COVID-19 infections in the summer. We divided the population into three sub-populations: long-term care (LTC) residents, university students, and the general population (everyone other than LTC residents and university students). We evaluated COVID-19 health outcomes in the city between August 15 and December 31 (4.5 months) with and without the introduction of 20,000 university students on September 1. We explored how the COVD-19 risk and prevention behaviours of the general population and the student community affect the incremental COVID-19 burden attributable to the arrival of the student population. Under different scenarios of community physical distancing effort and routine testing in students, we calculated the number of infected individuals, peak hospital resource demand, and number of deaths over time. Institutional ethics review was not required for this modeling study as human subjects were not involved.

A schematic of the model is presented in **Figure 1**. In the model, susceptible individuals may become infected through interaction with infected individuals who may or may not be aware of their infection status. Infection has a pre-symptomatic phase in which an infected individual can transmit the infection to others.^4,5,29^ Individuals may become aware of their infection status through symptom-based surveillance, contact tracing, or routine testing of asymptomatic and mildly symptomatic individuals. Individuals aware of their infection status with mild or moderate symptoms isolate at home to reduce disease transmission. Some patients develop severe symptoms requiring hospitalization or critical symptoms requiring mechanical ventilation (MV) in an intensive care unit or renal replacement therapy (RRT). Patients receive medically indicated care, unless resource demand exceeds capacity. When hospital capacity for a medically indicated resource has been reached, patients receive the next-best available care.

**FIGURE 1.**
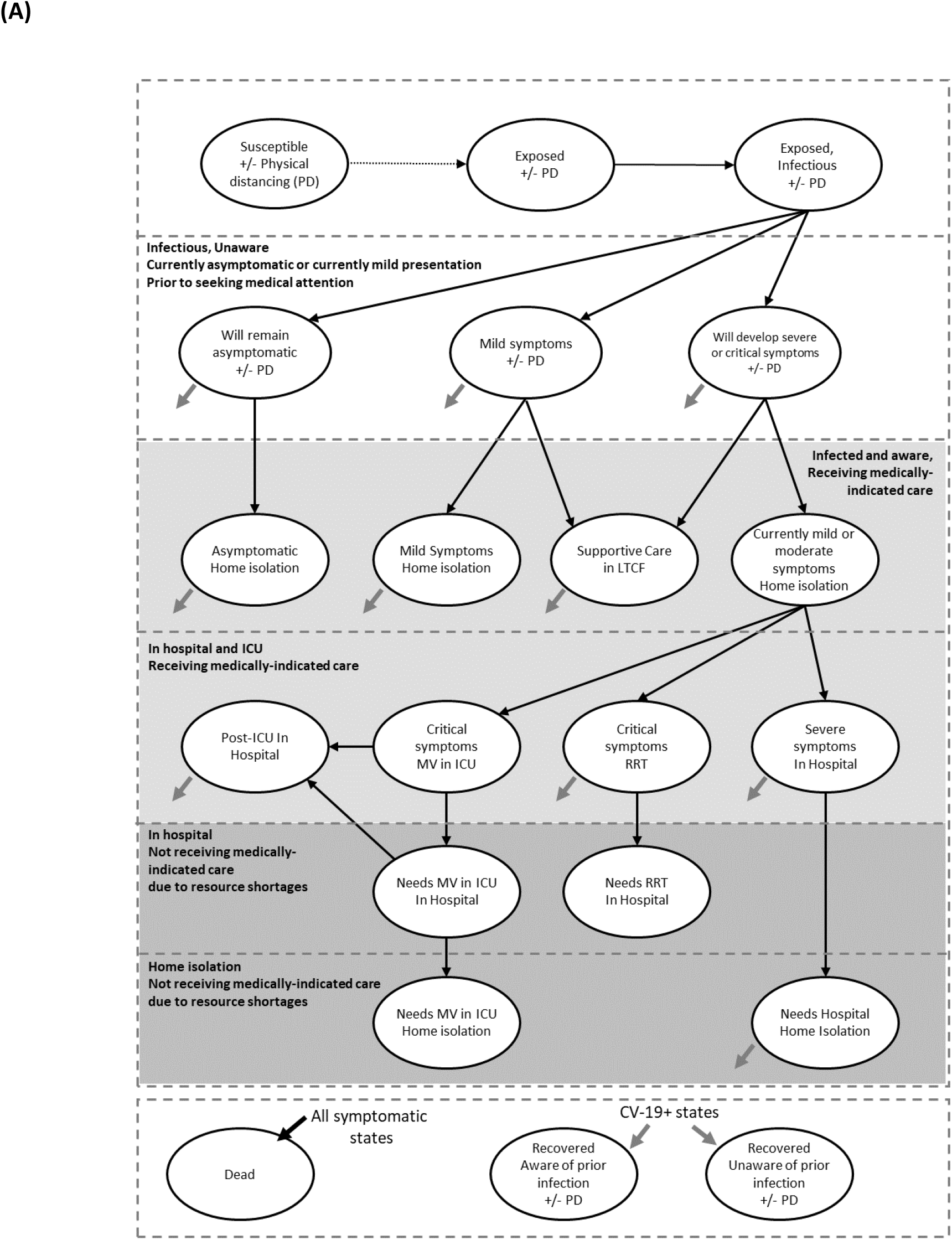
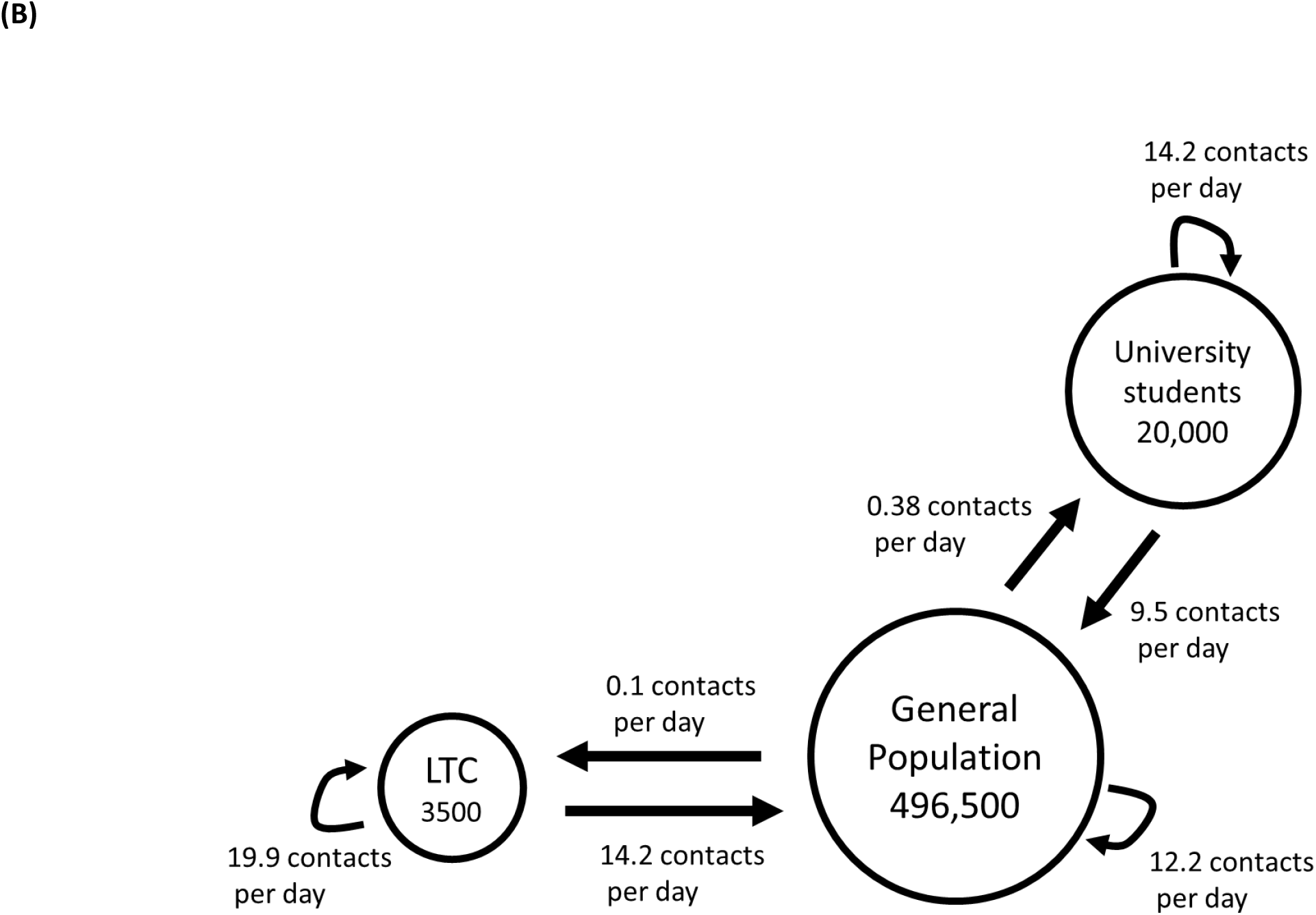
Model schematics of (A) COVID-19 health states and (B) close contact interactions between population subgroups. The number of contacts between groups indicated on the schematic represent the average number of contacts per day in a pre-COVID-19 era.

We estimated model parameters, including the duration of time spent in each health state, the infectiousness of COVID-19, demand for hospital resources and disease mortality conditional on disease severity, and the effectiveness of COVID-19 prevention strategies using the peer-reviewed literature, pre-published reports, and expert opinion (**Table 1**). We calibrated uncertain model inputs to the observed hospitalization and mortality outcomes in London, Ontario, Canada, a mid-size city with a large university population, between March 1 to August 15. Full details about the model structure and input parameter estimates are presented in the **Supplemental Methods**.

**TABLE 1.**
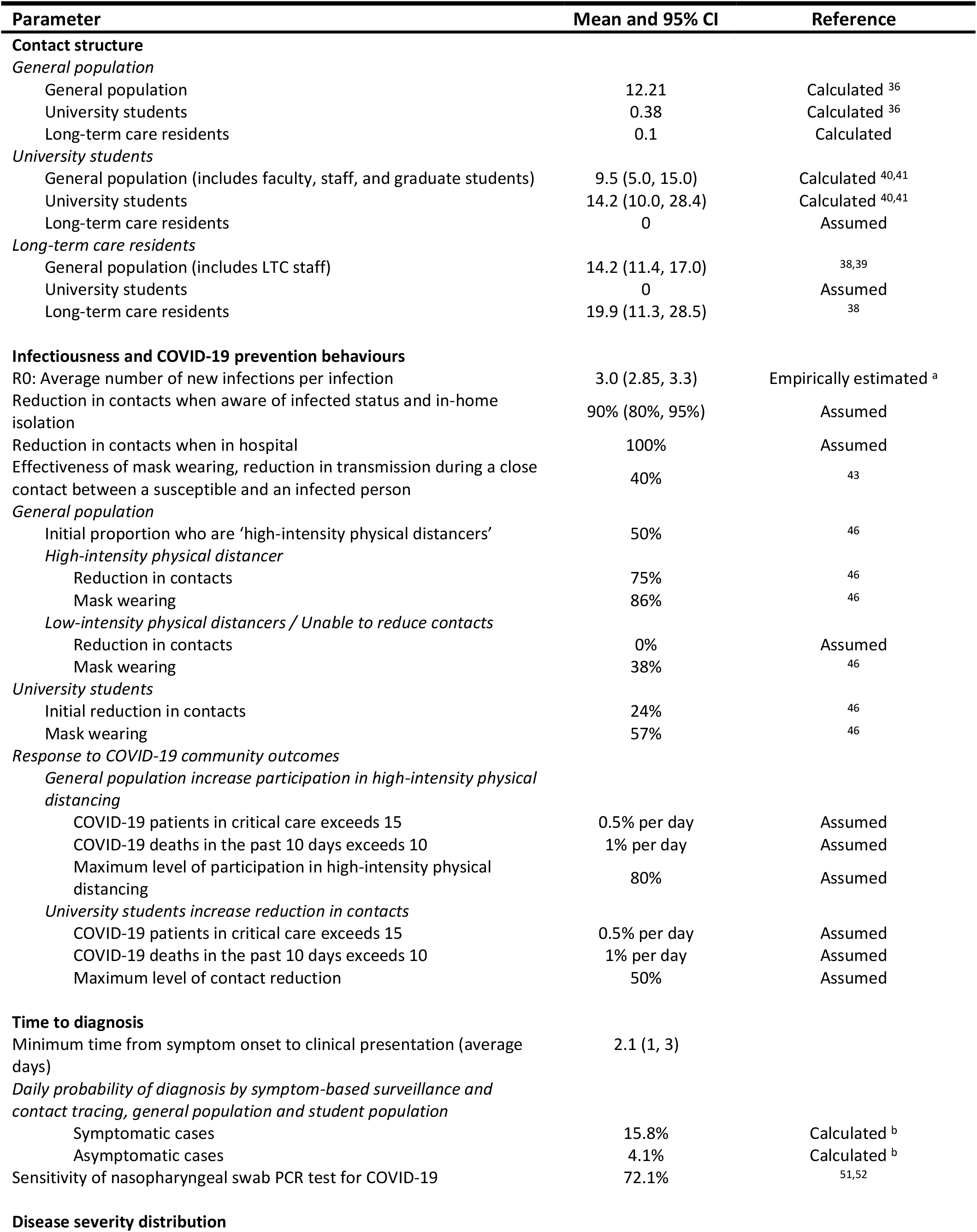
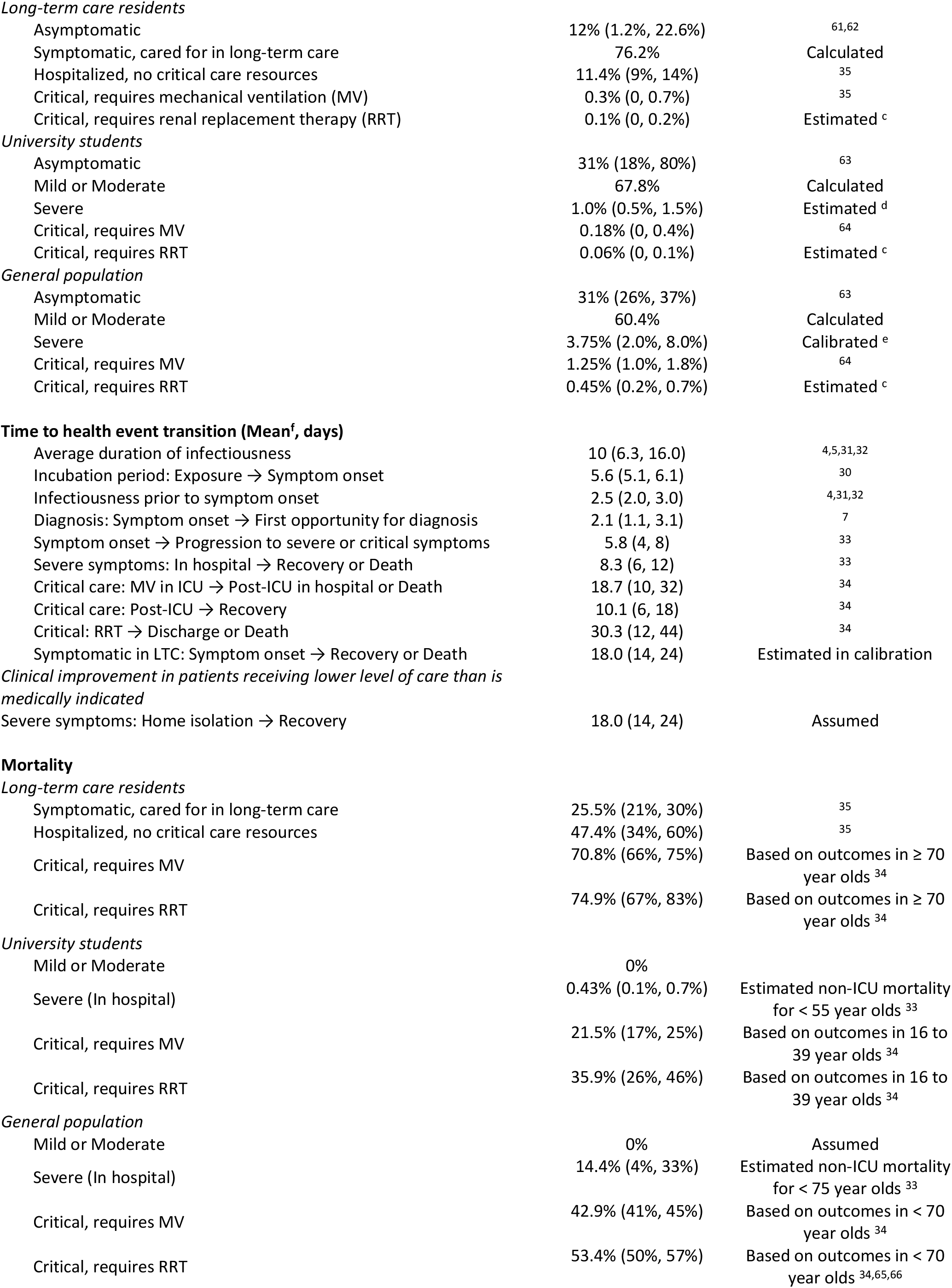
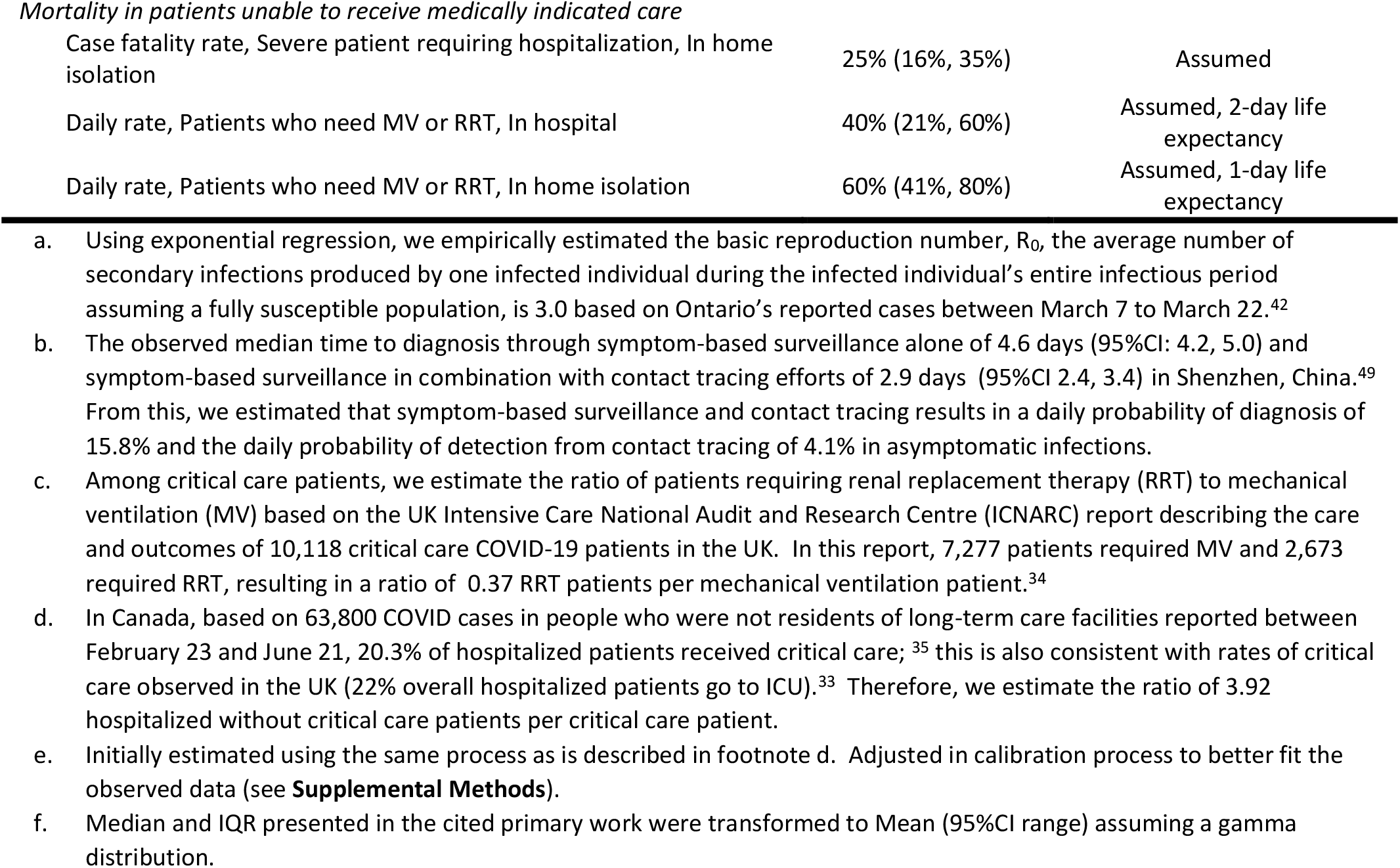
Base case parameters and sources. Mean and 95% confidence interval representing the uncertainty in the mean used in sensitivity analysis.

| Parameter | Mean and 95% CI | Reference |
| --- | --- | --- |
| **Contact structure** |  |  |
| *General population* |  |  |
| General population | 12.21 | Calculated <sup>36</sup> |
| University students | 0.38 | Calculated <sup>36</sup> |
| Long-term care residents | 0.1 | Calculated |
| *University students* |  |  |
| General population (includes faculty, staff, and graduate students) | 9.5 (5.0, 15.0) | Calculated <sup>40,41</sup> |
| University students | 14.2 (10.0, 28.4) | Calculated <sup>40,41</sup> |
| Long-term care residents | 0 | Assumed |
| *Long-term care residents* |  |  |
| General population (includes LTC staff) | 14.2 (11.4, 17.0) | <sup>38,39</sup> |
| University students | 0 | Assumed |
| Long-term care residents | 19.9 (11.3, 28.5) | <sup>38</sup> |
| **Infectiousness and COVID-19 prevention behaviours** |  |  |
| R0: Average number of new infections per infection | 3.0 (2.85, 3.3) | Empirically estimated <sup>a</sup> |
| Reduction in contacts when aware of infected status and in-home isolation | 90% (80%, 95%) | Assumed |
| Reduction in contacts when in hospital | 100% | Assumed |
| Effectiveness of mask wearing, reduction in transmission during a close contact between a susceptible and an infected person | 40% | <sup>43</sup> |
| *General population* |  |  |
| Initial proportion who are 'high-intensity physical distancers' | 50% | <sup>46</sup> |
| *High-intensity physical distancer* |  |  |
| Reduction in contacts | 75% | <sup>46</sup> |
| Mask wearing | 86% | <sup>46</sup> |
| *Low-intensity physical distancers / Unable to reduce contacts* |  |  |
| Reduction in contacts | 0% | Assumed |
| Mask wearing | 38% | <sup>46</sup> |
| *University students* |  |  |
| Initial reduction in contacts | 24% | <sup>46</sup> |
| Mask wearing | 57% | <sup>46</sup> |
| *Response to COVID-19 community outcomes* |  |  |
| *General population increase participation in high-intensity physical distancing* |  |  |
| COVID-19 patients in critical care exceeds 15 | 0.5% per day | Assumed |
| COVID-19 deaths in the past 10 days exceeds 10 | 1% per day | Assumed |
| Maximum level of participation in high-intensity physical distancing | 80% | Assumed |
| *University students increase reduction in contacts* |  |  |
| COVID-19 patients in critical care exceeds 15 | 0.5% per day | Assumed |
| COVID-19 deaths in the past 10 days exceeds 10 | 1% per day | Assumed |
| Maximum level of contact reduction | 50% | Assumed |
| **Time to diagnosis** |  |  |
| Minimum time from symptom onset to clinical presentation (average days) | 2.1 (1, 3) |  |
| *Daily probability of diagnosis by symptom-based surveillance and contact tracing, general population and student population* |  |  |
| Symptomatic cases | 15.8% | Calculated <sup>b</sup> |
| Asymptomatic cases | 4.1% | Calculated <sup>b</sup> |
| Sensitivity of nasopharyngeal swab PCR test for COVID-19 | 72.1% | <sup>51,52</sup> |
| **Disease severity distribution** |  |  |
| *Long-term care residents* |  |  |
| Asymptomatic | 12% (1.2%, 22.6%) | 61,62 |
| Symptomatic, cared for in long-term care | 76.2% | Calculated |
| Hospitalized, no critical care resources | 11.4% (9%, 14%) | 35 |
| Critical, requires mechanical ventilation (MV) | 0.3% (0, 0.7%) | 35 |
| Critical, requires renal replacement therapy (RRT) | 0.1% (0, 0.2%) | Estimated <sup>c</sup> |
| *University students* |  |  |
| Asymptomatic | 31% (18%, 80%) | 63 |
| Mild or Moderate | 67.8% | Calculated |
| Severe | 1.0% (0.5%, 1.5%) | Estimated <sup>d</sup> |
| Critical, requires MV | 0.18% (0, 0.4%) | 64 |
| Critical, requires RRT | 0.06% (0, 0.1%) | Estimated <sup>c</sup> |
| *General population* |  |  |
| Asymptomatic | 31% (26%, 37%) | 63 |
| Mild or Moderate | 60.4% | Calculated |
| Severe | 3.75% (2.0%, 8.0%) | Calibrated <sup>e</sup> |
| Critical, requires MV | 1.25% (1.0%, 1.8%) | 64 |
| Critical, requires RRT | 0.45% (0.2%, 0.7%) | Estimated <sup>c</sup> |
| **Time to health event transition (Mean<sup>f</sup>, days)** |  |  |
| Average duration of infectiousness | 10 (6.3, 16.0) | 4,5,31,32 |
| Incubation period: Exposure → Symptom onset | 5.6 (5.1, 6.1) | 30 |
| Infectiousness prior to symptom onset | 2.5 (2.0, 3.0) | 4,31,32 |
| Diagnosis: Symptom onset → First opportunity for diagnosis | 2.1 (1.1, 3.1) | 7 |
| Symptom onset → Progression to severe or critical symptoms | 5.8 (4, 8) | 33 |
| Severe symptoms: In hospital → Recovery or Death | 8.3 (6, 12) | 33 |
| Critical care: MV in ICU → Post-ICU in hospital or Death | 18.7 (10, 32) | 34 |
| Critical care: Post-ICU → Recovery | 10.1 (6, 18) | 34 |
| Critical: RRT → Discharge or Death | 30.3 (12, 44) | 34 |
| Symptomatic in LTC: Symptom onset → Recovery or Death | 18.0 (14, 24) | Estimated in calibration |
| *Clinical improvement in patients receiving lower level of care than is medically indicated* |  |  |
| Severe symptoms: Home isolation → Recovery | 18.0 (14, 24) | Assumed |
| **Mortality** |  |  |
| *Long-term care residents* |  |  |
| Symptomatic, cared for in long-term care | 25.5% (21%, 30%) | 35 |
| Hospitalized, no critical care resources | 47.4% (34%, 60%) | 35 |
| Critical, requires MV | 70.8% (66%, 75%) | Based on outcomes in ≥ 70 year olds <sup>34</sup> |
| Critical, requires RRT | 74.9% (67%, 83%) | Based on outcomes in ≥ 70 year olds <sup>34</sup> |
| *University students* |  |  |
| Mild or Moderate | 0% |  |
| Severe (In hospital) | 0.43% (0.1%, 0.7%) | Estimated non-ICU mortality for < 55 year olds <sup>33</sup> |
| Critical, requires MV | 21.5% (17%, 25%) | Based on outcomes in 16 to 39 year olds <sup>34</sup> |
| Critical, requires RRT | 35.9% (26%, 46%) | Based on outcomes in 16 to 39 year olds <sup>34</sup> |
| *General population* |  |  |
| Mild or Moderate | 0% | Assumed |
| Severe (In hospital) | 14.4% (4%, 33%) | Estimated non-ICU mortality for < 75 year olds <sup>33</sup> |
| Critical, requires MV | 42.9% (41%, 45%) | Based on outcomes in < 70 year olds <sup>34</sup> |
| Critical, requires RRT | 53.4% (50%, 57%) | Based on outcomes in < 70 year olds <sup>34,65,66</sup> |
| *Mortality in patients unable to receive medically indicated care* |  |  |
| Case fatality rate, Severe patient requiring hospitalization, In home isolation | 25% (16%, 35%) | Assumed |
| Daily rate, Patients who need MV or RRT, In hospital | 40% (21%, 60%) | Assumed, 2-day life expectancy |
| Daily rate, Patients who need MV or RRT, In home isolation | 60% (41%, 80%) | Assumed, 1-day life expectancy |

### POPULATION

We first establish the epidemic starting conditions in the city on August 15 before the potential arrival of students for fall semester. Based on calibration to COVID-19 outcomes in London, ON, at the start of the simulation, 40 individuals in the general population had active COVID-19 infections, 13.8 were exposed but not yet infectious, and 2,476 individuals had already recovered; thus, 493,937 individuals were susceptible at the beginning of the simulation. In sensitivity analysis, we vary the number of active COVID-19 infections in the general population at the start of the simulation.

In our analysis, 0.7% (3,500 individuals) of the population live in long-term care (LTC). Based on the model calibration we estimated that 75 LTC residents were recovered as of August 15. We assumed that there were no active infections in LTC residents on August 15.

We assume that 20,000 university students arrive on September 1. In the base case, we conservatively assume that no students are infected with COVID-19 upon arrival to campus. We vary this assumption in sensitivity analysis.

### CLINICAL OUTCOMES AND CLINICAL RESOURCE USE

A schematic of the health states and transition times for infected individuals is presented in **Appendix Figure 1**. We assumed a mean incubation period, the time from exposure to symptom-onset, of 5.6 days (observed median of 5.1 days [95%CI 2.2, 11.5]).^30^ In total, we estimated the average duration of infectiousness in asymptomatic and mild or moderate infections to be 10 days, including 2.5 days prior to symptom onset in individuals who do become symptomatic.^4,31,32^ We assumed that patients with severe and critical symptoms remain infectious until recovered, but that patient isolation protocols prevent transmission for those admitted to hospital.

We estimated hospital length of stay and mortality based on a report of over 20,000 hospitalized patients in the UK.^33^ We estimated length of stay and mortality for patients receiving critical care using the UK Intensive Care National Audit and Research Centre report describing the care and outcomes of 10,118 critical care COVID-19 patients.^34^ For LTC residents who are and are not hospitalized, we estimated the case fatality rate to be 47.4% and 25.5%, respectively, based on the observed outcomes in 680 Canadian LTC COVID-19 patients.^35^ Combining the assumptions about disease severity and severity-specific mortality rates, the overall infection fatality rate in our model was 25.1% for LTC residents, 1.3% for the general population, and 0.06% for university students. Among hospitalized cases, the fatality rate was 24.2% for the general population, and 5.2% for university students.

We did not consider mortality from causes other than COVID-19 in the model.

### CONTACT STRUCTURE

#### General population

Based on an extrapolation of the 2008 POLYMOD study in Europe to reflect network structure of the Canadian population, the average number of contacts per person in Canada is 12.6 per day, of which 1.0 contacts is aged 20-24.^36^ Assuming that the university student population adds 20,000 individuals aged 20-24 to a community with an otherwise typical Canadian age distribution, university students would comprise 38% of the population of people aged 20 to 24 in the community. As a result, we assume that a person in the ‘general population’ has contact with 0.38 university students per day. We calculated the average number of LTC contacts by calculating the number that would balance the staff and visitor contacts estimated for LTC residents, resulting in 0.1 LTC contacts per person in the general population.

#### Long-term care residents

We estimated that there are 19.9 resident-resident contacts per day (95% CI: 11.3 – 28.5) and 13.7 resident-staff contacts per day (95%CI: 11.4, 15.9) using a Canadian study of long-term care residents and staff (personal communication: S. Moghadas).^37,38^ This study did not capture resident-visitor contacts; we assumed 0.48 visitors per day based on the distribution of visit frequency in the 2012 Ohio Nursing Home Family Satisfaction Survey.^39^ Thus, in total, LTC residents have 14.2 contacts with the general population each day. We assumed no direct contact with university students.

#### University students

In our base case analysis, we assume that students have 23.7 contacts per day, based on a study at the University of Antwerp,^40^ and that 60% of those contacts are with other university students based on the age distribution of student’s reported contacts,^40,41^ with the remainder being with members of the ‘general population’ which includes staff and faculty of the university. Because studies evaluating university student contact patterns often occur during late fall and winter months, the contact patterns identified may not be fully representative of student contact rates at other times of the year. In sensitivity analysis we explored higher rates of student-student contacts in the first few weeks of the semester.

### INFECTION RISK AND DISEASE TRANSMISSION

Using exponential regression, we empirically estimated a basic reproduction number, R0, of 3.0 in the general population based on Ontario’s reported cases between March 7 to March 22.^42^ Using an average duration of infectiousness of 10 days and an average number of close contacts per person of 12.6,^36^ we calculate the probability of transmission between a susceptible and an infected person in close contact, in the absence of any interventions, to be 0.024.

Interventions such as physical distancing, which reduces the average number of contacts between susceptible and infected people, and mask wearing, which reduces the probability of disease transmission between contacts, can reduce the expected number of infections. We assume the effectiveness of cloth masks in reducing disease transmission is 40% based on a German study evaluating the effectiveness of real-world mask use.^43^ For people who are aware of their infection status and in home isolation, we assume a 90% reduction in contacts, which is at the high end of observed adherence to quarantine instructions in past epidemics.^44,45^

We subdivided the general population into two groups based on their intensity of COVID-19 prevention behaviours. Based on behaviours reported in an Angus Reid poll of Canadians, taken in the first week of August,^46^ we estimated that ‘high-intensity physical distancers’, representing 50% of the general population initially, reduce their average number of contacts by 75% (from 12.6 to 3.2 contacts per day) and that 86% of their remaining contacts are protected by a cloth mask. We assumed that the remainder of the population are not reducing their contacts, but are using a cloth mask to protect 38% of those contacts.^46^ In the base case, we assumed that university students initially reduce their contacts by 24% (from 23.7 to 18.0 contacts), which approximates a 75% reduction in contacts for the 32% of 18-24 year-olds who reported substantial COVID-19 prevention effort in the Angus Reid poll (32% × 75% = 24% reduction). In this same survey, 57% of 18-24 year-olds reported wearing a mask indoors with people outside their household and we use this as the base case level of mask wearing in the university student population.^46^

#### Responsive behaviour triggers

We assumed that the general population and university students respond to COVID-19 outcomes in the community. In practice, this response may be voluntarily adopted due to public concern over reported increases in COVID-19 cases, hospitalizations, and/or deaths or imposed through policies that re-establish the social and economic restrictions utilized in the earlier phases of the pandemic. We included two triggers that would cause both the general population and university students to increase their adoption of protective behaviors: a high level of COVID-19 patients in critical care and a high number of COVID-19 deaths.

The critical care trigger was set based on critical care capacity. The overall critical care capacity in Ontario is 14.2 beds per 100,000 population.^47^ Thus, in a city of 500,000, normal critical care capacity would be about 70 beds. While additional capacity can be created by seconding resources and personnel from other hospital services,^48^ based on expert opinion, substantial reductions in the provision of other types of health care (such as cancelling elective surgeries) would need to be undertaken if more than 30 critical care beds were used by COVID-19 patients. Therefore, we set one of the responsive behaviour triggers to activate when there are 15 COVID-19 patients in critical care, representing 50% of the capacity available to COVID-19 patients without modifying access to other health care services.

In the base case, the proportion of the general population who are ‘high intensity physical distancers’ increases by 0.5% each day if the number of COVID-19 patients in critical care exceeds 15 and by an additional 1.0% each day if the number of COVID-19 deaths in the past 10 days exceeds 10, up to a maximum of 80% participation in high-intensity physical distancing. Similarly, we assume students’ reduction in contacts increases at the same rate in response to the same triggers, but up to a maximum of a 50% reduction in contacts (23.7 contacts to 11.9 contacts).

### DIAGNOSIS BY SYMPTOM-BASED SURVEILLANCE, CONTACT TRACING, AND ROUTINE TESTING

For the general population and university students, we assumed the minimum time from symptom onset to diagnosis to be 2.1 days, which is consistent with the minimum time needed to self-assess, seek medical attention, and receive diagnostic results.^7^ The observed median time to diagnosis through symptom-based surveillance alone was 4.6 days (95%CI: 4.2, 5.0) and symptom-based surveillance in combination with contact tracing efforts was 2.9 days (95%CI 2.4, 3.4) in Shenzhen, China.^49^ From this, we estimated that symptom-based surveillance and contact tracing results in a daily probability of diagnosis of 15.8% for symptomatic infections and a daily probability of detection (from contact tracing) of 4.1% for asymptomatic infections. This combination of assumptions resulted in approximately 22% of infected individuals being identified, consistent with the overall rates of diagnosis implied by preliminary serology data in Ontario.^50^

We considered policy alternatives of routine screening for COVID-19 in university students at various screening frequencies, including every 14, 10, 7, 5, and 3 days. We also considered the value of a one-time universal screening three weeks after student arrival. We identified the date of the one-time testing by identifying the date that minimized the number of total infections over the semester. We assumed that testing will be performed with the standard COVID-19 nasopharyngeal swab followed by PCR analysis with a test sensitivity of 72.1%.^51,52^

## RESULTS

### City epidemic outcomes without the introduction of the university student population

Without the introduction of the student population, the base case assumptions for the general population and LTC residents leads to a total of 3,900 infections over 4.5 months (August 15 to December 31). In this scenario, infections, hospitalizations, and daily deaths do not peak until the new year (**Appendix Table 1**). Demand for critical care (mechanical ventilation or renal replacement therapy) peaks at 27 beds early in the new year, and a total of 31 COVID-19 deaths occur between August 15 and December 31 (60 deaths by January 31, 2021). The timing and magnitude of the city’s COVID-19 outbreak, excluding any impacts from students, is determined by the initial number of COVID-19 infections in the community, the level of participation in physical distancing, the responsiveness of the community to increasing critical care cases and COVID-19 deaths, and the proportion of contacts that are protected with mask wearing (**Appendix Figures 5-7**).

### Effect of introducing 20,000 university students to the community

In the base case, we conservatively assumed that students would bring no undiagnosed infections of COVID-19 to the community and would immediately engage in physical distancing efforts that resulted in a similar average contact reduction to the general population (reduction of 24%, from average of 23.7 contacts to 18.0 contacts). Even so, university students continue to have a higher average number of contacts than members of the general population. As a result, in this base case scenario, the introduction of students to the community increases the total number of infections by 3,399 infections, representing an 87% increase (from 3,900 to 7,299) (**Figure 2**). Only 28% (960) of these incremental infections occur in the student population (representing 4.8% of students becoming infected). Of the remaining 72% of incremental infections, 2,428 occur in the general population (0.5% of the susceptible general population), and 11 occur in long-term care (0.3% of the susceptible LTC population). The larger absolute increase in infections in the general population occurs due to the connectivity of the university community with the general population and the relative size of the general population. The increase in infections among LTC residents, despite our assumption that there are no direct contacts between university students and LTC residents, occurs due to the increase in infections in the general population. The higher number of infections results in an increase in hospitalizations, demand for critical care, and COVID-19 deaths. Peak critical care occupancy increases by 48% (from 27 to 40 beds). These outcomes include the mitigation effects of the responsive behaviour of the community to seeing high levels of COVID-19 hospitalizations. The introduction of students to the community also moves up the timing of responsive behaviours, with the threshold of 15 COVID-19 patients in critical care being reached 3 weeks earlier (**Appendix Table 2**).

**TABLE 2.** Infections averted in the general population with 5-day testing and one-time testing of students compared to a policy of no routine asymptomatic testing (symptom-based surveillance and contact tracing only). Scenarios vary the proportion of infections in the student population that are asymptomatic and timing and level of students contact reductions. We calculate the expected number of critical care admissions averted and COVID-19 deaths averted to be 1.7% and 1.32% of general population infections averted which includes hospitalizations and deaths which may occur after December 31 to all individuals infected prior to December 31.

| Scenario | Without students | With students | 5-day testing compared to no routine testing |  |  |  | One-time testing three weeks after student arrival compared to no routine testing |  |  |  |
| --- | --- | --- | --- | --- | --- | --- | --- | --- | --- | --- |
|  |  |  | Infections averted | % averted in the general pop'n | Critical care adm'n averted | COVID-19 deaths averted | Infections averted | % averted in the general pop'n | Critical care adm'n averted | COVID-19 deaths averted |
| 31% asymptomatic in students |  |  |  |  |  |  |  |  |  |  |
| Base case physical distancing (24% reduction in contacts immediately) | 3,900 | 7,299 | 1,392 | 70% | 16.5 | 12.8 | 127 | 81% | 1.8 | 1.4 |
| Two weeks of 2.0x activity among students, followed by 24% reduction in contacts | 3,900 | 10,294 | 2,661 | 64% | 29.1 | 22.6 | 361 | 66% | 4.0 | 3.1 |
| Low level of physical distancing (15% reduction in contacts immediately) | 3,900 | 11,209 | 3,448 | 61% | 35.9 | 27.9 | 91 | 64% | 1.0 | 0.8 |
| Two weeks of 2.0x activity among students, followed by 24% reduction in contacts; 60% of general population engaged in high-intensity physical distancing | 543 | 7,317 | 3,737 | 65% | 41.0 | 31.8 | 337 | 66% | 3.8 | 2.9 |
| 50% asymptomatic in students |  |  |  |  |  |  |  |  |  |  |
| Base case physical distancing (24% reduction in contacts immediately) | 3,900 | 7,445 | 1,533 | 70% | 18.2 | 14.2 | 175 | 81% | 2.4 | 1.9 |
| Two weeks of 2.0x activity among students, followed by 24% reduction in contacts | 3,900 | 10,582 | 2,931 | 65% | 32.4 | 25.1 | 414 | 67% | 4.7 | 3.7 |
| 80% asymptomatic in students |  |  |  |  |  |  |  |  |  |  |
| Base case physical distancing (24% reduction in contacts immediately) | 3,900 | 7,524 | 1,603 | 69% | 18.8 | 14.6 | 40 | 68% | 0.5 | 0.4 |
| Two weeks of 2.0x activity among students, followed by 24% reduction in contacts | 3,900 | 10,889 | 3,209 | 65% | 35.3 | 27.4 | 337 | 60% | 3.4 | 2.7 |

**FIGURE 2.**
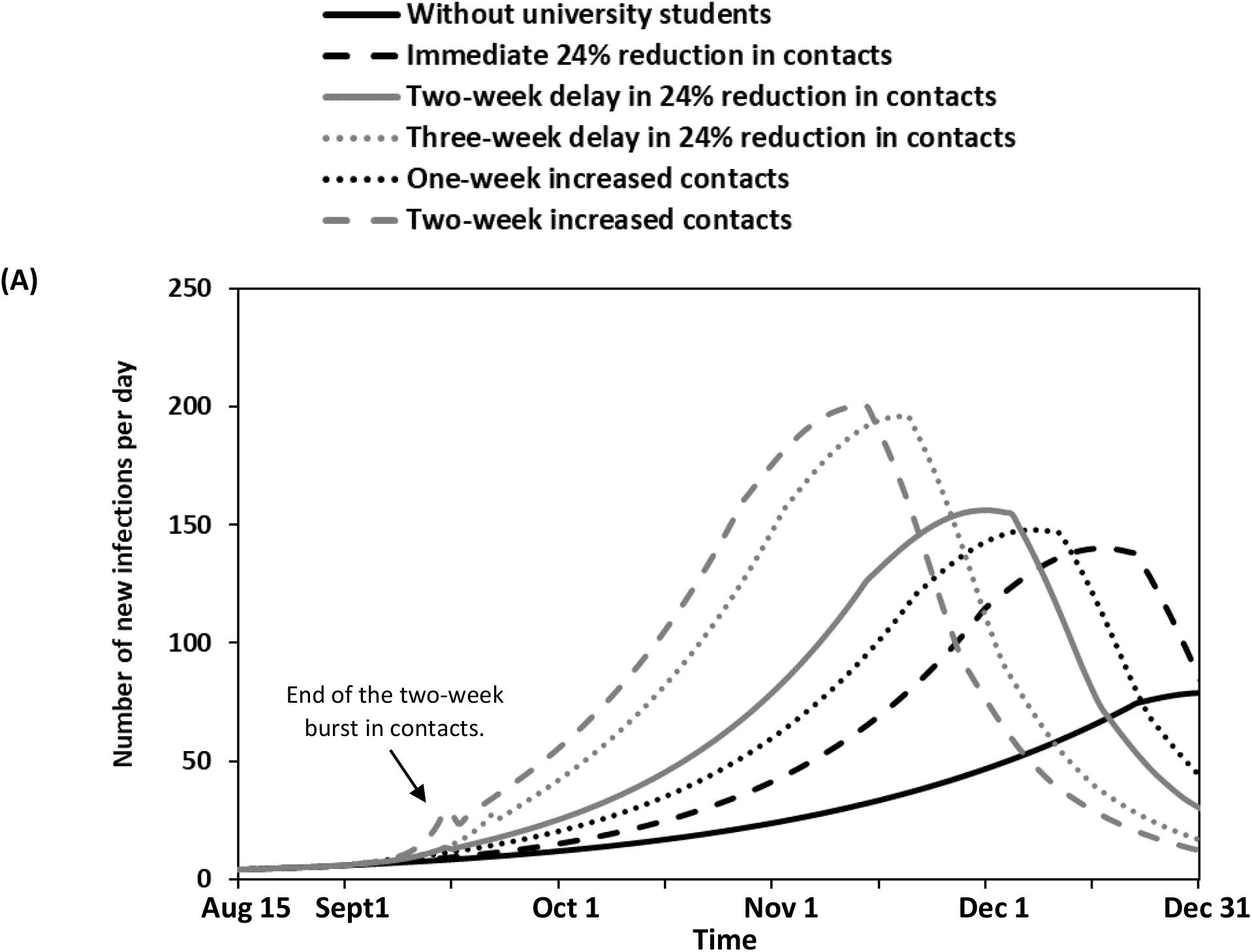
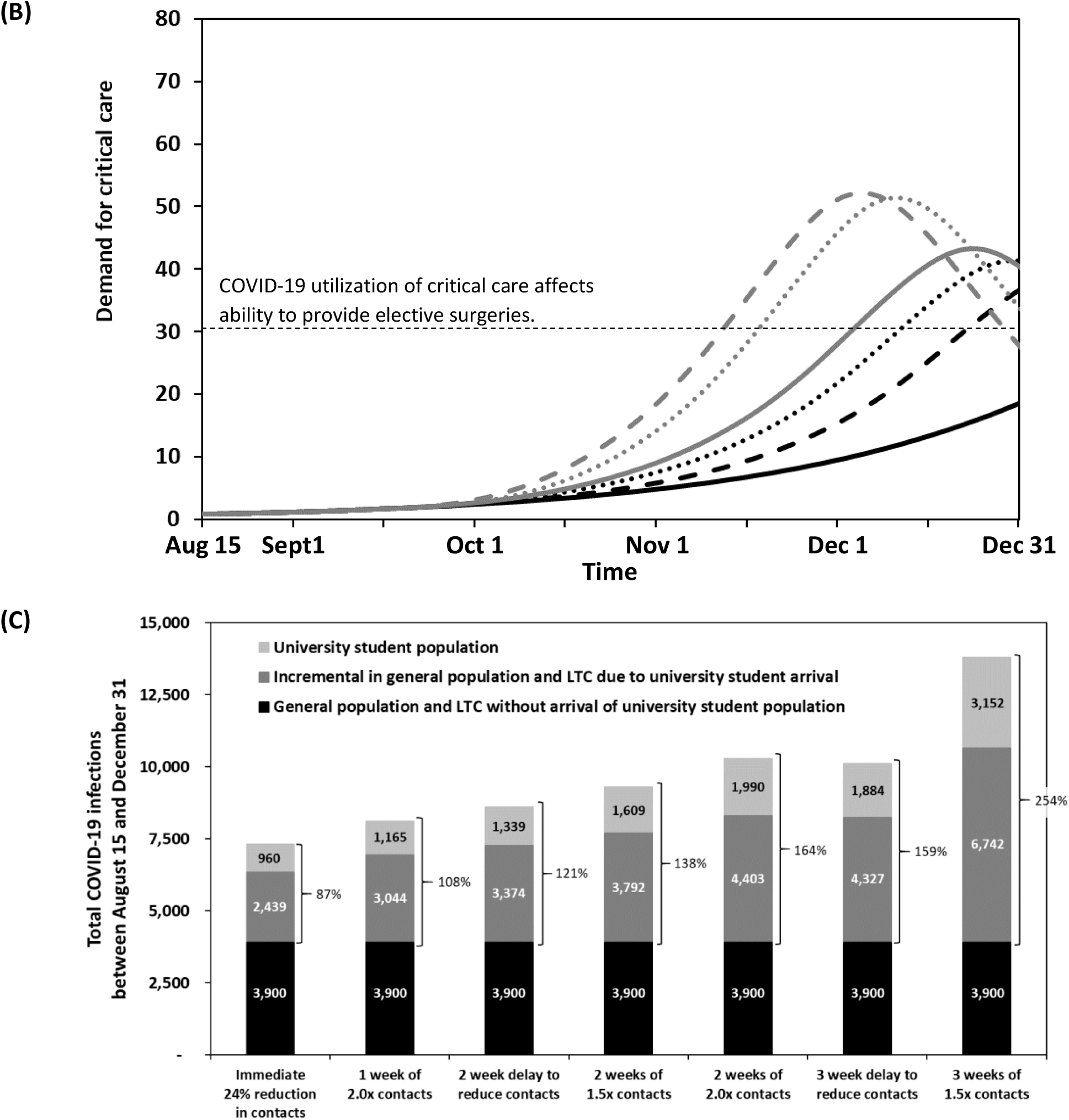
Epidemic outcomes in a city of 500,000 with and without the introduction of 20,000 university students on September 1. Scenarios consider different initial physical distancing behaviours in the university student population. (A) Number of new infections per day; (B) the number of people medically indicated for critical care each day; and, (C) the cumulative number of COVID-19 infections between August 15 and December 31. Numerical results are provided in **Appendix Table 4**.

If some students arrive exposed or asymptomatically infected, the total number of infections occurring over the course of the semester increases. For example, if 10 students arrive infected, the number of infections increases by 1,047 over the base case. The impact of students arriving already exposed or infected in the community is most substantial on the timing of peak infections, peak hospitalizations, peak critical care utilization, and the timing of responsive behaviour triggers. Compared to the scenario without the introduction of the student population, responsive behaviours are triggered 6.5 weeks earlier if 10 students are infected when they arrive (**Appendix Table 3**).

### Initial level of student contact behaviour

Our estimates of average student contacts per day were based on surveys completed at times other than during the first few weeks of the school year, which normally involve a large number of social activities. Even if muted, the first few weeks of the new academic year may still result in substantially more contacts than a typical pre-COVID era day. Therefore, we explored the consequences associated with students having an increased number of student contacts for one or two weeks upon arrival to the community. For example, if students simply delay implementing physical distancing for two weeks, the total number of infections attributable to the introduction of the students into the community increases by 4,713 infections (from 3,900 to 8,613 infections). In this scenario, the arrival of students would result in a 120% increase in the number of infections the community. As in the base case, the majority of these incremental infections occur in the general population (**Figure 2**).

If students have twice the pre-COVID era number of contacts with other students for two weeks (37.9 contacts per day, 28.4 of whom are other students), the total number of infections in the community increases by 6,394 infections representing a 164% increase in the number of infections the community would expect without the students and leading to an additional 72 COVID-19 deaths (**Appendix Figure 8, Appendix Table 4**). Delays in the implementation of contact reductions, or short-term increases in the number of student-student contacts increases demand for critical care resources and shortens the time until critical care beds dedicated to COVID-19 patients exceeds 30 beds (**Figure 2B**). The impact on total infections is mitigated by the earlier activation of responsive behaviour triggers, which occurs 8.3 weeks after the arrival of the students and 8 weeks earlier than without the arrival of the student population.

### Effectiveness of routine asymptomatic screening targeted at students

Because young people have a high rate of asymptomatic and mild presentation, routine testing of students has been proposed for university campus settings. Testing students every 28 days results in very little reduction in the number of infections but requires a large number of tests (714 students tested per day). Testing every 14 days, as is recommended for staff at long-term care facilities,^53^ reduces the number of infections in the student population from 960 to 735, a 23% reduction, and reduces the overall number of infections due to the introduction of the university student population by 22% (from 3,399 to 2,653) (**Figure 3**).

**FIGURE 3.**
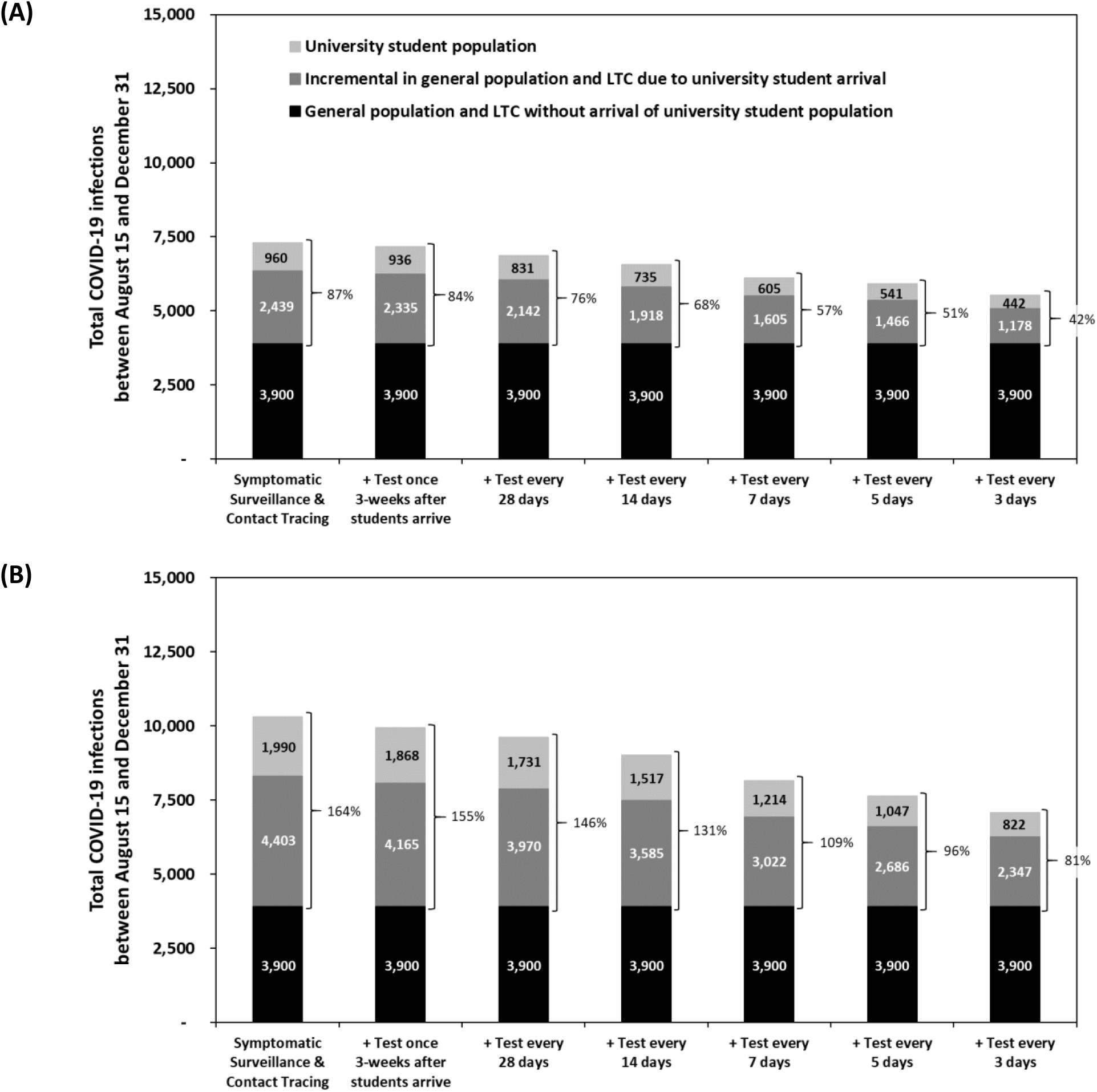
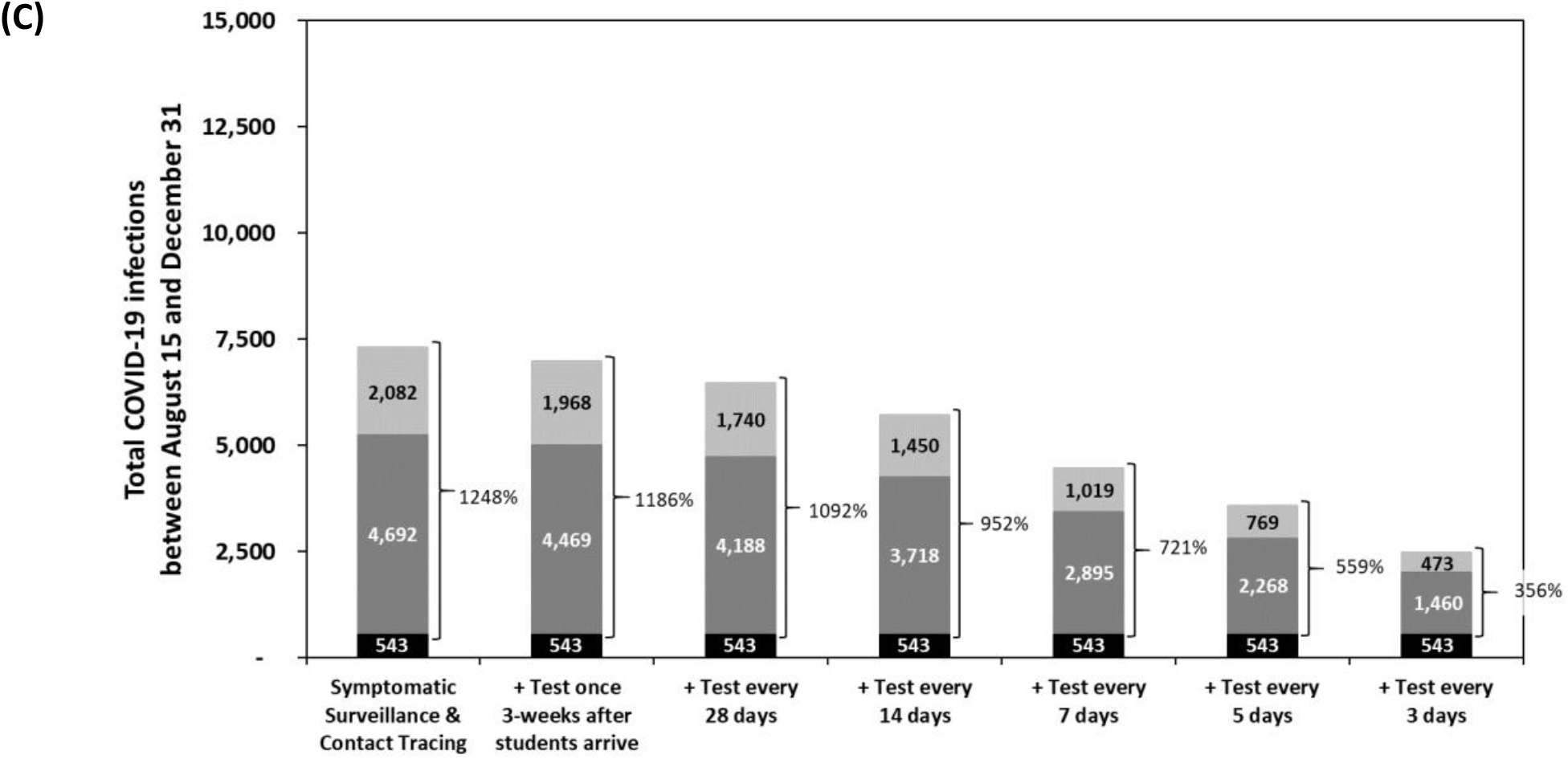
Cumulative number of COVID-19 infections between August 15 and December 31 in a city of 500,000 with and without the introduction of 20,000 university students on September 1. Scenarios in each panel differ in the frequency with which students undergo routine testing for COVID-19. In panel (A), students have an average 24% reduction in contacts compared to normal student social interaction behaviour (average of 23.7 contacts reduced to 18.0 contacts) immediately upon arrival with no short-term increase in contacts; in panel (B), students double their contacts with other students for the first two weeks and then implement a 24% reduction in their contacts; in panel (C), students double their contacts with other students for the first two weeks and then implement a 24% reduction in contacts and 60% of the general population is participating in high-intensity physical distancing (compared to 50% in the base case and other scenarios presented in this figure). Other outcomes for these scenarios are reported in **Appendix Table 5**.

More frequent testing reduces infections further. Testing students every 5 days reduces the number of infections among the student population by 44% (from 960 to 541) and reduces the total number of infections due to the introduction of the university student population to the community by 41% (from 3,399 to 2,007). Routine testing has greater impact in the scenarios when students engage in a short-term increase in the number of contacts early in the term. In the scenario in which students double their student contacts for the first two weeks of term, testing every 5 days reduces the number of infections in the student population by 47% (from 1,990 to 1,047), reduces the total number of infections due to the introduction of the university student population by 42% (from 6,394 to 3,732), and delays the activation of responsive behaviour triggers by 1 week (**Appendix Table 5**).

Routine testing of students also averts critical care admissions and COVID-19 mortality in the general population, because approximately two-thirds of averted infections are prevented in the general population (**Table 2**). In the base case analysis, in which students immediately reduce their contacts compared to a pre-COVID-19, routine screening every 5 days averts 16.5 critical care admissions and 12.8 deaths. In scenarios in which students have short term increases in their contact behaviours or lower levels of contact reduction, routine testing every 5 days averts more than 29 critical care admissions and greater than 20 COVID-19 deaths.

Sensitivity analysis revealed that routine testing of university students was more valuable when students have a higher rate of asymptomatic infections (**Table 2**) and in scenarios in which the differences in transmission risk between the university students and the general population were greater. For example, in a scenario in which the city had a high level of engagement in physical distancing routine screening of the student population can avert a large number of infections because in these scenarios the city expects very little COVID-19 transmission without the introduction of the student population (**Figure 3C**). Conversely, in scenarios in which the city is engaged in a low level of physical distancing and so expects a large number of infections with or without the student population, the difference in risk profile between the city and the university populations decreases, as does the benefits of targeting prevention efforts at the university population.

### Effectiveness of one-time screening targeted at students

Routine testing to identify and isolate asymptomatic infections for the purposes of reducing community transmission risk requires a large number of tests each day and may strain community testing resources. We also evaluated the benefits of a one-time universal screening event occurring three weeks after the students arrive. Compared to routine testing every 5 days, which would require more than 400,000 tests to be performed over the semester, this strategy would only require 20,000 tests. Through the isolation of identified cases, one-time testing is able to immediately decrease the daily number of new infections in the student population and, so, indirectly in the general population (**Appendix Figure 9**). In the case that students double their contacts with other students for a period of two-weeks, this strategy prevents 361 infections (122 infections in students, 238 infections in the general population, and 1 infection in LTC residents), 4.0 critical care admissions, and 3.1 COVID-19 deaths (**Table 2**). One-time screening does not significantly impact the timing of peak infections, resource utilization, or the time that responsive behaviour triggers are activated (**Appendix Table 5**).

### Sensitivity analysis

We performed extensive sensitivity analysis exploring the impact of general population and student population COVID-19 prevention behaviours on the incremental impact of introducing students into the community.

The negative impacts of introducing the student population can be partially mitigated through high uptake of COVID-19 preventive behaviours in the student population including high rates of contact reduction or if the rate of mask wearing significantly exceeds the level reported by 18 to 24 year-olds (**Appendix Figure 10, Appendix Table 6**). For example, if students immediately reduce their contacts by, on average, 30% (from 23.7 to 16.6 contacts per day) and wear masks to protect 65% of their remaining contacts, the incremental number of infections attributable to the arrival of the student population can be reduced to 1,698 infections (from 3,900 to 5,598), representing only a 44% increase over the number of infections the community would expect without the students, and delays the activation of responsive behaviour triggers by 1 week.

The magnitude of the impact of the introducing the student population is also determined by the COVID-19 prevention behaviours of the general population. Counter-intuitively, the relative impact of introducing the student population is greatest when the prevention efforts by the general population are high (**Appendix Figure 11**). For example, when 60% of the general population are participating in high-intensity physical distancing, without students the number of new infections per day is nearly constant over time, resulting in a very low level of cumulative infections over the semester (total of 543 infections). Introduction of the students results in 939 additional infections, more than doubling the total number of infections expected in the city without the addition of the student population (**Appendix Table 2**). In such a scenario, because the student population is an important determinant of city outcomes, the impact of routine COVID-19 screening in the student population is greater (**Figure 3C**).

## DISCUSSION

In this analysis, we consider the COVID-19 impacts of the re-opening of a destination university in a mid-sized city with varying epidemiological contexts. Without the return of university students, we devised several scenarios involving moderate to severe fall COVID-19 waves based on difference levels of preventive behaviours in the general population. The return of a large number of students always worsens these waves, even under the conservative assumptions that arriving students do not introduce any new infections to a community and that they immediately adopt substantial COVID-19 prevention behaviours. This is because university students have nearly double the number of contacts as the general population due to residing in shared or congregate living situations, working in the service sector, and higher levels of social activity. In the scenarios we considered, this increase in infections was substantial, potentially doubling of the total number of COVID-19 infections in the city over the fall. Notably, we found that more than two-thirds of the incremental infections attributable to the arrival of the student population occurred in the general population and, as a result, substantially impacted the city’s health care resource needs, COVID-19 mortality in the community, and accelerated the need for responsive behaviour which may be in the form of re-engaged social and economic restrictions.

Our base case finding that the return of university students would increase the number of infections by 87% is likely conservative. In this analysis, we assumed that no students arrive already infected, students do not engage in short term increases in contacts upon arrival, and that students respond to adverse community COVID-19 outcomes by increasing the intensity of their prevention behaviours at the same rate as the general population. At the very least, it may take time for students to fully adopt protective measures; moving into dorms, orientation events, and start of semester social events (even if not officially sanctioned by the university) may result in higher-than-normal levels of contact for at least the first few weeks. In the analyses in which we consider short-term increases in the average number of student-student contacts, we show that a higher level of contact for just the first one or two weeks can dramatically increase the total number of infections experienced by the city over the semester.

Our analysis found that the majority of the increase in infections due to the arrival of students occurred in the general population, not in the student population itself. While university campuses may seem like isolated bubbles, the general community and university students are intertwined, as staff and faculty interact with students on campus and students interact with others off-campus in work, living, and social settings. Previous studies modeling university populations did not account for infections in the broader off-campus community.^20-24^ However, we have shown that including the general population when modeling COVID-19 transmission on university campuses is critical, since this population bears the brunt of the incremental morbidity and mortality burden of COVID-19. As a result, university policies that either discourage student return to the community,^54^ such as offering coursework fully online, or mitigate COVID-19 risks for students returning to campus, such as screening for COVID-19 symptoms and routine COVID-19 testing in asymptomatic individuals, can have substantial impacts on the city’s COVID-19 burden. Imposing restrictions on students’ off-campus social behavior may be practically difficult, necessitating modified behaviors in the general population in response to university re-opening, such as additional reductions in social contacts to balance the increased risk of returning university students. For example, in the base case, the increase in infections due to student arrival could be mitigated if the proportion of the general population engaged in high-intensity physical distancing increased by 4.6% (from 50% to 54.6%). This illustrates the idea of “risk budgets”, where increased risk in one domain of a community necessitates reducing risk in another to keep COVID-19 impacts below desired thresholds.^55^

Our analysis indicates that routine testing of all students every 5 days can avert a substantial number of infections, critical care admissions, and COVID-19 deaths. In the base case, we estimate that testing every 5 days prevents 16.5 critical care admissions and 12.8 deaths and, in the scenario in which students double their contacts with other students for two weeks, we estimate that testing every 5 days prevents 29.1 critical care admissions and 22.6 deaths. Using the simplifying assumption that all deaths averted will be in 60-year-olds and a willingness to pay of $50,000 per life-year-gained, the economic value of these deaths averted is $13.4 million to $23.6 million, translating to a value of $33 to $59 per test. This calculation underestimates the benefits of testing by not accounting for savings due to averted critical care admissions and the community economic benefits of delaying social and economic restrictions; despite this, because we estimate the lab cost of nasopharyngeal testing for COVID-19 at $80 per test at our center, high-frequency routine testing is likely only cost effective using batch testing strategies.

Alternatively, one-time universal testing of students after an initial burst of social activity among students may be more feasible operationally and economically. We estimated that this strategy can prevent 360 infections, 4.0 critical care admissions, and 3.1 COVID-19 deaths corresponding to an approximate economic value of $3.3 million or $164 per test. This strategy is most effective at changing the trajectory of new infections if testing occurs after a short-term period of high social activity and is less effective if students have consistent but lower levels of contact reductions (e.g., immediately reducing their contacts, but by only 15% instead of the 24% in the base case).

An important limitation of our analysis is that we assume students with a COVID-19 diagnosis will be willing and able to self-isolate effectively. However, it may be challenging for students to isolate from roommates or refrain from using shared facilities, like bathrooms and kitchens, without dedicated university-organized isolation facilities.^56,57^ Furthermore, adherence to isolation guidance may be low, especially if the majority of infections in university students are asymptomatic or mild. During the H1N1 pandemic, a survey of symptomatic university students found that only 41% of students followed recommendations to stay home until well.^16^ In the base case, we also assume that students are equally responsive as the general population to COVID-19 outcomes in the community reducing their contacts in response to high numbers of critical care hospitalizations and deaths. In reality, university students may be less aware of the impacts of COVID-19 on hospital resources and less concerned about COVID-19 generally given their lower risk of adverse outcomes. The extent and speed with university students respond to hospitalizations and deaths in the local community will impact the number of infections experienced by the community and the benefits of routine testing in the student population.

Compared to other modeling studies of COVID-19 on university campuses, the total number of infections and the number of infections averted by testing we estimate over the semester are modest. This is because we assume that both university students and the general population will increase their self-protective behavior (physical distancing) in response to high numbers of COVID-19 hospitalizations and deaths, either through individual decision-making or adaptive community policies. These adaptive behaviors are more realistic than assuming a population will maintain the same behavior no matter the severity of local COVID-19 conditions. Thus, in our analysis, testing is being layered onto a robust and reactive mitigation response.

Our model includes only three sub-populations and so does not include many other aspects of age-structured mixing or age-dependant health outcomes. The model does not estimate the impact of COVID-19 patient utilization on the provision or effectiveness of other health care services and the model does not account for death from causes other than COVID-19.^58^ The model includes community transmission by stratified random mixing but does not include additional imported index cases from other cities, which may occur into the general population or the student population, nor does the model include the stochastic consequences of super-spreading events. Especially early on in an epidemic or when cases have been brought to very low levels, dynamics are sensitive to random outcomes in the number of new infections resulting from each case (e.g., ‘Patient 31’ in South Korea^59^ and ‘Patient One’ in Italy^60^).

## Conclusion

We developed a model-based analysis to estimate the impact of a relatively large student population on the COVID-19 outcomes of a mid-sized city with relatively few cases of COVID-19 prior to the return of students. Our analysis is relevant to a number of mid-sized cities in North America with relatively large university and college populations. Because university students have substantially more contacts than the general population, due to congregate living environments, high-density social activities, and disproportionate employment in the service sector, the introduction of university students substantially increases the number of COVID-19 infections and decreases the time until responsive behaviours are activated. Substantial uncertainty exists in the level of contact reduction that students will choose, or is feasible given their living, transit, and work situations. Public health interventions, such as routine testing, targeted at this population prevents infections in the entire population, improving community health related and unrelated to COVID-19.

## Data Availability

Input data are largely publicly available and references with links are provided. Contact authors for any additional information.

## Acknowledgements

The authors are grateful to Dr. Seyed Moghadas, Professor of Applied Mathematics and Computational Epidemiology at York University, Toronto, Ontario for sharing data relating to the contact patterns of long-term care residents.

